# Infants are more susceptible to COVID-19 than children

**DOI:** 10.1101/2021.05.02.21256474

**Authors:** Char Leung

## Abstract

Angiotensin-converting enzyme 2 (ACE2) has been found to mediate the host cell entry of SARS-CoV-2 that causes COVID-19. However, the link between ACE2 and the observed susceptibility of SARS-CoV-2 infection remains elusive. In contrast, observational studies can help identify the susceptibility biomarker of SARS-CoV-2 infection, those associated with age for example. Data of all PCR tests performed in the state of São Paulo of Brazil were gathered from the government database and were analyzed using multivariate logistic regression. Adjusted odds ratios for positive test results were calculated with the adjustment of age, gender, and comorbidities. Over 1.7 million test results were included in the study of which 38% were positive. Elderly was most vulnerable to SARS-CoV-2 infection. While underages were less susceptible than adults aged below 60 years, susceptibility was not equal among different pediatric groups. It is found that age and susceptibility to SARS-CoV-2 infection are not inversely related, but U-shaped, with infants more susceptible than children. Biomarkers that are linearly associated with age cannot explain the reduced susceptibility in children. These include lymphocyte count and cross-reactive antibodies against other coronaviruses that offers cross-protection. The expression level of ACE2 may still be able to explain but further investigations are needed.

## Introduction

Severe acute respiratory syndrome coronavirus 2 (SARS-CoV-2) is an enveloped, positive-sense, and single-stranded RNA virus, one of the seven coronaviruses pathogenic to human, causing coronavirus disease 2019 (COVID-19) that originated in Wuhan of China. Its crown like appearance is given by the spike glycoprotein that is expressed on the surface of the virus particle. The entry process involves binding of the spike glycoprotein to the cellular receptor angiotensin-converting enzyme 2 (ACE2) which mediates viral attachment and spike protein priming by type II transmembrane serine protease (TMPRSS2) allowing fusion of viral and cellular membranes [1].

ACE2 is abundantly expressed on the airway epithelial cells, causing a range of respiratory infection symptoms such as cough, nasal congestion, and dyspnea as well as fatal complications including respiratory failure and sepsis. Consequently, a large body of research has been devoted to study the expression and distribution of ACE2 in human.

However, the link between ACE2 and the observed susceptibility of SARS-CoV-2 infection remains elusive for a number of reasons. First, the expression level of ACE2 in different cohorts remains debated, that between males and females for example [2], despite males more vulnerable to SARS-CoV-2 infection. Of note, ACE2 also mediates the host cell entry of SARS-CoV but lower prevalence of SARS in males was observed in Taiwan, Hong Kong, and Singapore [3], weakening the association between ACE2 expression level and susceptibility. Second, the role ACE2 plays in SARS-CoV-2 infection is not clear. While it mediates host cell entry, there is also evidence suggesting a protective effect of ACE2 in pneumocytes [4]. ACE2 is a counter-regulator against ACE by converting angiotensin II to angiotensin_(1-7)_ through hydrolysis [5] and angiotensin_(1-7)_ promotes vasodilation, protecting against lung inflammation and fibrosis [6,7]. In addition, the lower expression of ACE2 means an upregulation of angiotensin II, that has been found to induce the production of IL-6 [8], a proinflammatory cytokine associated the increased risk for mortality in patients infected with SARS-CoV-2 [9].

Therefore, the expression level of ACE2 alone is not sufficient to determine the susceptibility to SARS-CoV-2 infection. On the other side of the coin, observational studies can help identify and verify potential susceptibility biomarkers. For example, reduced susceptibility to symptomatic infection with SARS-CoV-2 has been observed in children [10] and lymphocyte count that decreases with age may play a role in susceptibility. Nevertheless, it remains uncertain whether neonates are less susceptible to SARS-CoV-2 infection because of their distinctive immune systems.

Against this background, the present work describes the age-dependent susceptibility to SARS-CoV-2 infection, including the unequal susceptibility among different pediatric groups, so as to shed light on the possible age-varying susceptibility biomarkers, using the publicly available data of all PCR tests performed in the state of São Paulo of Brazil.

## Methods

Data were gathered on the 29th November 2020 from a publicly available online system managed by the Ministry of Health of the Brazilian Government, allowing certified healthcare professionals to register suspected and confirmed cases of COVID-19 for surveillance purposes. Therefore, the database does not only include symptomatic patients but also asymptomatic ones as well as those who had epidemiological links to confirmed cases. Because it is a nationwide reporting system, only basic demographic and medical measures, including results of diagnostic tests for SARS-CoV-2 infection. Furthermore, the nature of self-reporting means that the validity of these measures depends on the judgement and discretion of the certified medical professionals.

Due to the size of the dataset, only cases in the state of São Paulo were considered. Furthermore, only patients with conclusive PCR-test (nasopharyngeal swab) results were included in the study as it remains the gold standard for COVID-19 diagnosis. Patients with age unidentified were excluded from the study. The following data were abstracted: (i) gender, (ii) age at PCR-test taken, (iii) comorbidities, (iv) asymptomaticity and (v) PCR-test result. Patients were divided into 7 different age group with the corresponding definition in parentheses: neonate (aged 30 days or below), infant (aged above 30 days but below 2 years), young child (aged 2 years or above but below 6 years), child (aged 6 years or above but below 12 years), adolescent (aged 12 years or above but below 18 years), adult (aged 18 years or above but below 60 years), and elderly (aged 60 years or above).

To see the effect of age on SARS-CoV-2 infection, a multivariate logistic regression model was used to estimate the age-dependent odds ratios for PCR positive results, adjusted for confounding with gender, comorbidities, that may affect the result of the PCR tests, and asymptomaticity to reduce possible selection bias. In addition, statistical comparisons were made between PCR-positive and -negative group using appropriate statistical tests depending on the type of data and normality condition. Mann-Whitney tests or t-tests were used for continuous variables whereas Fishers exact tests were used for dichotomous variables.

## Results

A total of 2,012,765 suspected and confirmed cases with the use of PCR test were found in the database, including all cases dated from February to November 2020. Of these, 1,765,415 cases had conclusive PCR test results. After the removal of cases failing to meet the inclusion criteria, a total of 1,721,403 cases were eligible for inclusion of which 38% were positive for SARS-CoV-2 infection.

Results of statistical tests comparing the baseline characteristics between groups are shown in Table 1. Due to the very large sample, all measures were found significant. Furthermore, all measures were selected in the logistic regression. Some comorbidities such as diabetes, chronic heart and kidney diseases were identified as independent risk factors for SARS-CoV-2 infection. In contrast, immunodeficiency, chronic respiratory disease, obesity, pregnancy and chromosomal disease were associated with decreased risk of SARS-CoV-2 infection. Interestingly, all these odds ratios consistently lied between 0.7 and 1.2. In line with existing studies [2], males were found more susceptible to infection.

**Table 1.** Summary of the studied cohort, a comparison between PCR-positive and -negative group

|  | PCR-positive (n) | PCR-negative (n) | p (difference between PCR+ vs PCR-) | Adjusted OR (95% CI) |
| --- | --- | --- | --- | --- |
| Baseline characteristics, % unless otherwise stated |  |  |  |  |
| Age, years | 39.4 (658946) | 36.9 (1062457) | < 0.001 |  |
| Neonate | 0.045 (299/658946) | 0.1 (633/1062457) | < 0.001 | 0.763 (0.664,0.876) |
| Infant | 0.5 (3377/658946) | 1.2 (13082/1062457) | < 0.001 | 0.399 (0.385,0.415) |
| Young child | 0.6 (4166/658946) | 1.4 (15302/1062457) | < 0.001 | 0.427 (0.412,0.442) |
| Child | 1.1 (7313/658946) | 1.9 (20693/1062457) | < 0.001 | 0.562 (0.547,0.577) |
| Adolescent | 2.4 (15872/658946) | 3.0 (32260/1062457) | < 0.001 | 0.787 (0.772,0.803) |
| Adult | 82.3 (542373/658946) | 81.3 (864079/1062457) | < 0.001 | 1 (reference) |
| Elderly | 13.0 (85546/658946) | 11.0 (116408/1062457) | < 0.001 | 1.157 (1.146,1.169) |
| Male | 45.5 (299733/658924) | 41.3 (438484/1062449) | < 0.001 | 1.217 (1.210,1.225) |
| Asymptomatic | 1.8 (11918/658946) | 6.0 (63551/1062457) | < 0.001 | 0.279 (0.273,0.284) |
| Comorbid conditions, % |  |  |  |  |
| Immunodeficiency | 0.8 (5549/658946) | 0.9 (9846/1062457) | <0.001 | 0.851 (0.823,0.880) |
| Diabetes | 5.0 (32643/658946) | 3.7 (39618/1062457) | <0.001 | 1.196 (1.177,1.215) |
| Chronic heart disease | 8.1 (53688/658946) | 6.8 (71777/1062457) | <0.001 | 1.074 (1.060,1.087) |
| Chronic kidney disease | 0.5 (3007/658946) | 0.4 (4046/1062457) | <0.001 | 1.071 (1.021,1.123) |
| Chronic respiratory disease | 3.0 (19596/658946) | 3.9 (41786/1062457) | <0.001 | 0.727 (0.714,0.739) |
| Obesity | 0.3 (1998/658946) | 0.4 (3951/1062457) | <0.001 | 0.771 (0.730,0.814) |
| Pregnant | 0.4 (2665/658946) | 0.5 (5738/1062457) | <0.001 | 0.792 (0.756,0.830) |
| Chromosomal disease | 0.3 (1890/658946) | 0.3 (3636/1062457) | <0.001 | 0.829 (0.784,0.877) |

For age, statistical tests comparing the two groups indicated significantly greater proportion of PCR-positive in adults and elderly. After the adjustment for asymptomaticity and comorbidities, however, susceptibility was not linear but U-shaped with neonates appearing to be significantly more vulnerable to SARS-CoV-2, than infants and young children, as shown by the adjusted odds ratios (ORs). In addition, all pediatric groups appeared less susceptible to adults. These ORs with the corresponding 95% confidence intervals are graphically shown in Figure 1.

**Figure 1.**
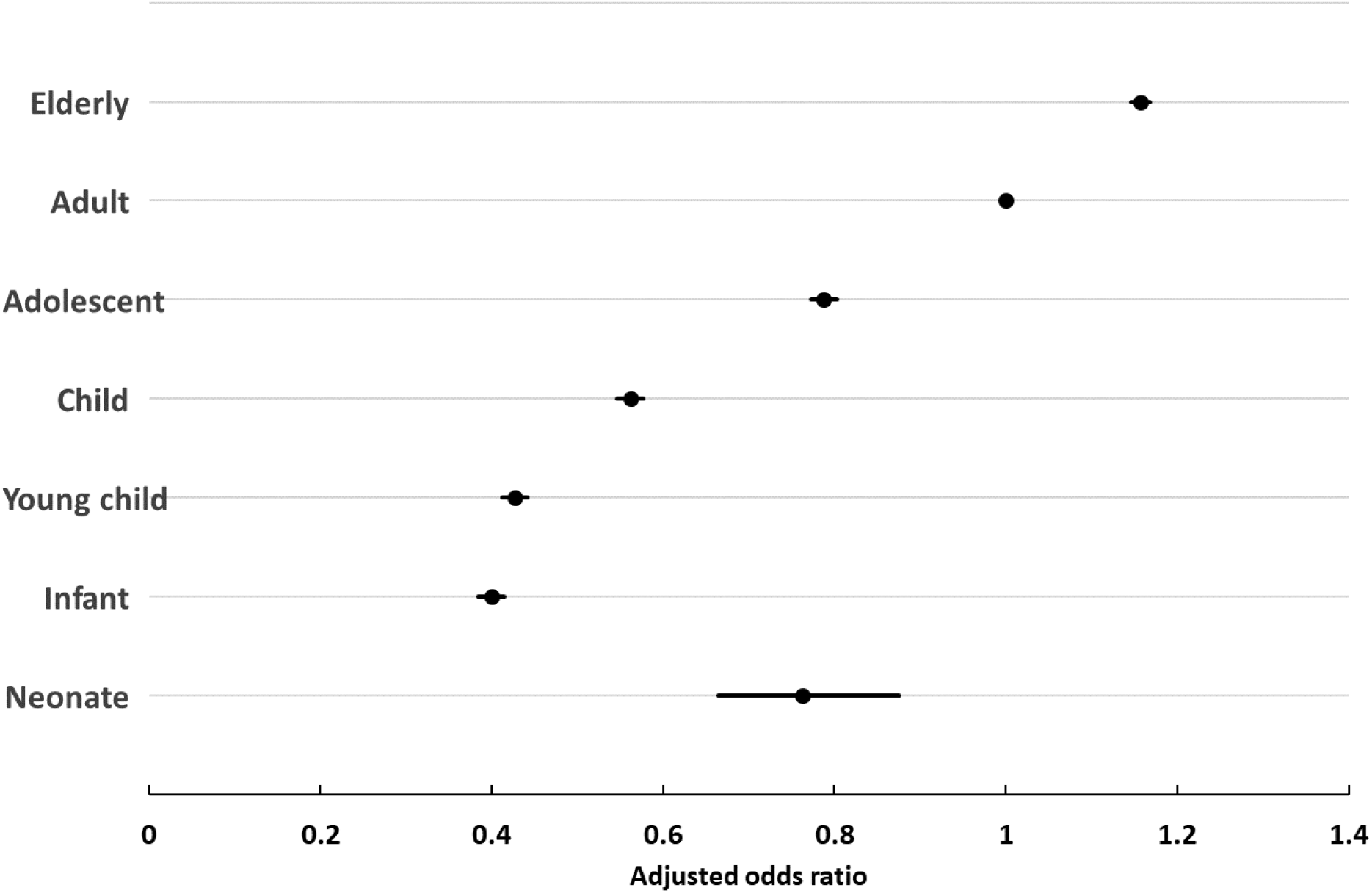
Age-dependent adjusted odds ratios with 95% CI (black lines)

## Discussion

Using the very large publicly available data collected in the state of São Paulo, the present work verifies that newborns, children and adolescents are less susceptible to SARS-CoV-2 infection, compared to adult, and that newborns are more susceptible than children. The relation between age and the susceptibility to SARS-CoV-2 infection appears U-shaped.

Increased susceptibility in neonates has previously been observed although no attention was paid. For instance, studies concerning more granular breakdown of the age distribution of confirmed cases have shown higher prevalence in those aged below 5 years [11-13] than those between 6 and 10. In contrast, lower prevalence was observed in those aged 10 years or below [14,15], compared with other age groups, suggestive of increased susceptibility in neonates.

The U-shaped relationship observed implies that biomarkers that are linearly associated with age alone cannot explain the reduced susceptibility in children, lymphocyte count for instance. If it is associated with the susceptibility to SARS-CoV-2 infection, the prevalence of COVID-19 should be lower in infants than in children because lymphocyte count decreases with age [16].

It has also been suggested by Wong et al [17] that previous exposure to other coronaviruses might provide cross-protection to children because seroconversion to NL63 and 229E, another two strains of coronavirus pathogenic to human, may produce antibodies to coronaviruses that have some degree of neutralizing and cross-protective activity against infection to another coronavirus. If this hypothesis is correct, an inverted U-shaped relation between seroprevalence and age should be observed because the present work suggested increased susceptibility in neonates. However, studies [18,19] cited by the authors rather found a U-shaped relation.

While the expression level of ACE2 in neonates remains unclear as discussed earlier, it is possible to hypothesize a U-shaped relation between ACE2 and age. Coupled with the finding of a high positive correlation between plasma ACE and ACE2 activity in healthy individuals [20], a U-shaped relation between age and ACE level with higher ACE activity in infants (aged between 0.5 and 2 years) than young children (aged between 2 to 4 years) [21] suggests a U-shaped relation between age and ACE2 level. Nevertheless, further investigation is needed to verify this hypothesis as well as the pathological role of ACE2.

Because of its role of membrane fusion, TMPRSS2 may also play a role in the susceptibility of SARS-CoV-2. A recent study found that the expression of TMPRSS2 increases with age and that infants and children expressed similarly low levels of TMPRSS2 [22]. In light of the observed U-shaped relation between age and susceptibility, it is suggested that TMPRSS2 alone is not sufficient to explain susceptibility.

Finally, the results also suggested the relation between susceptibility and different comorbidities. Immunodeficiency is suggested to be associated with reduced risk for SARS-CoV-2 infection. This is in line with a current review study and the authors proposed that weaker immune response determined a milder disease presentation and thus underdiagnosis [23]. Similarly, lower prevalence of asthma was observed in COVID-19 patients and type-2 cytokines have been proposed to play a role [24]. Unfortunately, little is known for other comorbidities. For instance, there is not yet clear evidence on the relation between obesity and susceptibility [25].

Selection bias is the major limitation of the study. For instance, symptomatic patients seeking medical care are more likely to be tested. However, the database also included asymptomatic patients that had epidemiologic links to confirmed cases and asymptomaticity was included in the statistical analysis to reduce the selection bias. Unless all individuals in São Paulo were tested, selection bias remains. In this regard, the studied cohort represented a significant proportion of the population to reduce the bias. With a population of 44 million in São Paulo and 1.72 million PCR tests performed, the test rate is approximately 3,910 per 100,000 inhabitants, about the same as Belgium and above Germany and France [26].

## Data Availability

Data are available upon reasonable requests.

## Abbreviation

PCR: polymerase chain reaction.

